# UV light dosage distribution over irregular respirator surfaces. Methods and implications for safety

**DOI:** 10.1101/2020.04.07.20057224

**Authors:** Aurora Baluja, Justo Arines, Ramón Vilanova, Julio Cortiñas, Carmen Bao, Maite Flores

**Author notes:** **Correspondence:** Aurora Baluja. Department of Anesthesiology, Intensive Care and Pain Management, Complexo Hospitalario Universitario. Santiago de Compostela. Instituto de Investigación Sanitaria (IDIS). Travesía da Choupana s/n, Santiago de Compostela, 15706 A Coruña, Spain.

## Abstract

**Background and Objectives:** The SARS-CoV-2 pandemic has led to a global decrease in personal protective equipment (PPE), especially filtering facepiece respirators (FFRs). Ultraviolet-C wavelength is a promising way of decontamination, however adequate dosimetry is needed to ensure balance between over and underexposed areas and provide reliable results. Our study demonstrates that UVGI light irradiance varies significantly on different respirator angles and propose a method to decontaminate several masks at once ensuring appropriate dosage in shaded zones.

**Methods:** An UVGI irradiator was built with internal dimensions of 69.5 × 55 × 33 cm with three 15W UV lamps. Inside, a grating of 58 × 41 × 15 cm was placed to hold the masks. Two different flat fold respirator models were used to assess irradiance, four of model Aura 9322 3M of dimensions 17 × 9 × 4cm (tri-fold), and two of model SAFE 231FFP3NR (bi-fold) with dimensions 17 × 6 × 5 cm. A spectrometer STN-SilverNova was employed to verify wavelength spectrum and surface irradiance. A simulation was performed to find the irradiance pattern inside the box and the six masks placed inside. These simulations were carried out using the software DIALUX EVO 8.2.

**Results:** The data obtained reveal that the irradiance received inside the manufactured UVGI-irradiator depends not only on the distance between the lamps plane and the base of the respirators but also on the orientation and shape of the masks. This point becomes relevant in order to assure that all the respirators inside the chamber receive the correct dosage.

**Conclusion:** Irradiance over FFR surfaces depend on several factors such as distance and angle of incidence of the light source. Careful irradiance measurement and simulation can ensure reliable dosage in the whole mask surface, balancing overexposure. Closed box systems might provide a more reliable, reproducible UVGI dosage than open settings.

## INTRODUCTION

The novel human coronavirus (SARS-CoV-2) pandemic has led to a global, critical decrease in personal protective equipment (PPE), especially filtering facepiece respirators (FFRs). Due to this shortage, multiple recommendations have arisen, in particular those related to the use of ultraviolet germicidal irradiation (UVGI, 254 nm) for decontamination (Heimbuch et al., 2011; Lindsley et al., 2015; Viscusi et al., 2011). As of 27/03/2020 CDC issued new guidelines to reuse masks (CDC, 2020) acknowledging that decontaminated N95 mask limited reuse may be necessary in dire shortage situations.

UVGI acts primarily over surfaces. Thus, surface shape, incidence angle and distance related to the light source are key factors for local irradiance. The resulting UV dose (fluence) is therefore the product of the irradiance by exposure time. Given the high spread potential and severity of SARS-CoV-2, local overdose may be sacrificed, in order to minimize contamination risk by underexposure, as most FFRs can tolerate higher than germicidal doses. However, protocols for mask decontamination inside rooms with powerful UV-C sources might not ensure an even irradiance distribution among masks placed at different angles from the lamp.

The main objective of this study is to demonstrate that UVGI light dosage varies significantly on different respirators depending of their position inside the disinfection chamber and propose a method to decontaminate several masks at once ensuring appropriate dosage in shaded zones.

## METHODS

### UVGI device

An UVGI box irradiator was built with internal dimensions of 69.5 cm length, 55 cm width, 33 cm height. Inside, a grating of 58×41×15 cm was placed in order to hold the masks. Three 15W lights HNS 15W G13 (OSRAM) were located at the upper limit in three of the four walls of the box. Each lamp provides 4.9 W in the UVC wavelength. The plane containing the three lamps is parallel to the bottom. The grating that will hold the respirators is placed over the bottom. The distance between the lamps plane and the grating was evaluated and measurements were taken to find the more homogeneous irradiance inside the UVGI chamber. The whole internal surface of the chamber was coated with a matte aluminum insulating lining. Aluminum is known to present a good reflection in the UV-C wavelength range (Bass et al., 2009; Welch et al., 2018). The matte finish improves light diffusion providing a safer, more even irradiance distribution -softer shadowing-, at the expense of increased irradiation times due to lower reflectivity.

### Irradiance measurement

A spectrometer STN-SilverNova, with a sensitivity range between 190 nm and 1110 nm (2nm resolution) equipped with a STN-CR2-cosine corrector was used to verify the 254 nm emission spectrum. The spectro-radiometer is radiometrically calibrated allowing us to measure the irradiance received inside the chamber and evaluate the several critical positions. In order to evaluate the optimal orientation of the respirators inside the chamber, several irradiance measurements were done.

Additionally, to evaluate if there is difference in the irradiance received by the respirators, when they are placed in different positions inside the chamber, as well as to evaluate the shadows in terms of irradiance when several respirators are disinfected at the same time, the following measurements were performed. Two different flat fold respirator models were used to assess irradiance, four of model Aura 9322 3M of dimensions 17 × 9 × 4cm (trifold), and two of model SAFE 231FFP3NR (bi-fold) with dimensions 17 × 6 × 5 cm. Both masks have different sizes and heights upon the grating.

### Irradiance simulation

Some simulations were also made, in order to find the shadows and areas with less irradiance inside the box. These simulations were carried out using the software DIALUX EVO v. 8.2 (DIAL GmbH, Lüdenscheid, Germany). This software is freely downloadable and extensively used in the industry of indoor and outdoor illumination. DIALUX offers only information in arbitrary units (AU) for any surface and volume.

Simulation provides a more general understanding of the distribution of light in arbitrary units (AU) on the chamber, possible shadows inside the chamber and relevance of the correct placement of the masks, in order to receive sufficient irradiation. Modeling can also help us determine irradiance distribution in relative units. Finally, modeling can guide further chamber development (optimal lamp placement) or optimize costs for new units (e.g lamp set energy consumption).

Two masks models were simulated: model SAFE 231FFP3NR and the model Aura 9322 3M. The first model was simulated with a truncated pyramid of dimensions (length, width, height) 17cm × 4cm × 9cm. The base of 4 cm corresponds to the case were the masks are slightly open. The second model was simulated with a truncated pyramid of dimensions 17cm × 9cm × 5cm. The masks were placed in two rows and three columns as they are planned to be in the disinfection box. Additionally, two different orientations were simulated with respect to the long side of the box, parallel and perpendicular.

### COVID-area setting

The irradiator, equipped with four wheels, was placed in the COVID ICU of a tertiary-care hospital (16 beds) in a separate room more than 2 m away from any COVID patient. The placement was inside the COVID area to avoid contamination elsewhere. The box was equipped with a main on-off switch and an interlock as a safety mechanism that turned the lamps on when closed and off upon lid opening.

## RESULTS

### Irradiance measurement

The spectrum of the lamp (see Figure 1) was measured, showing its peak at the 254 nm with a Full Width Half Maximum (FWHM) of 4.84 nm. A lamp heating time around 5 min was observed, in order to obtain a stable emission.

**Figure 1:**
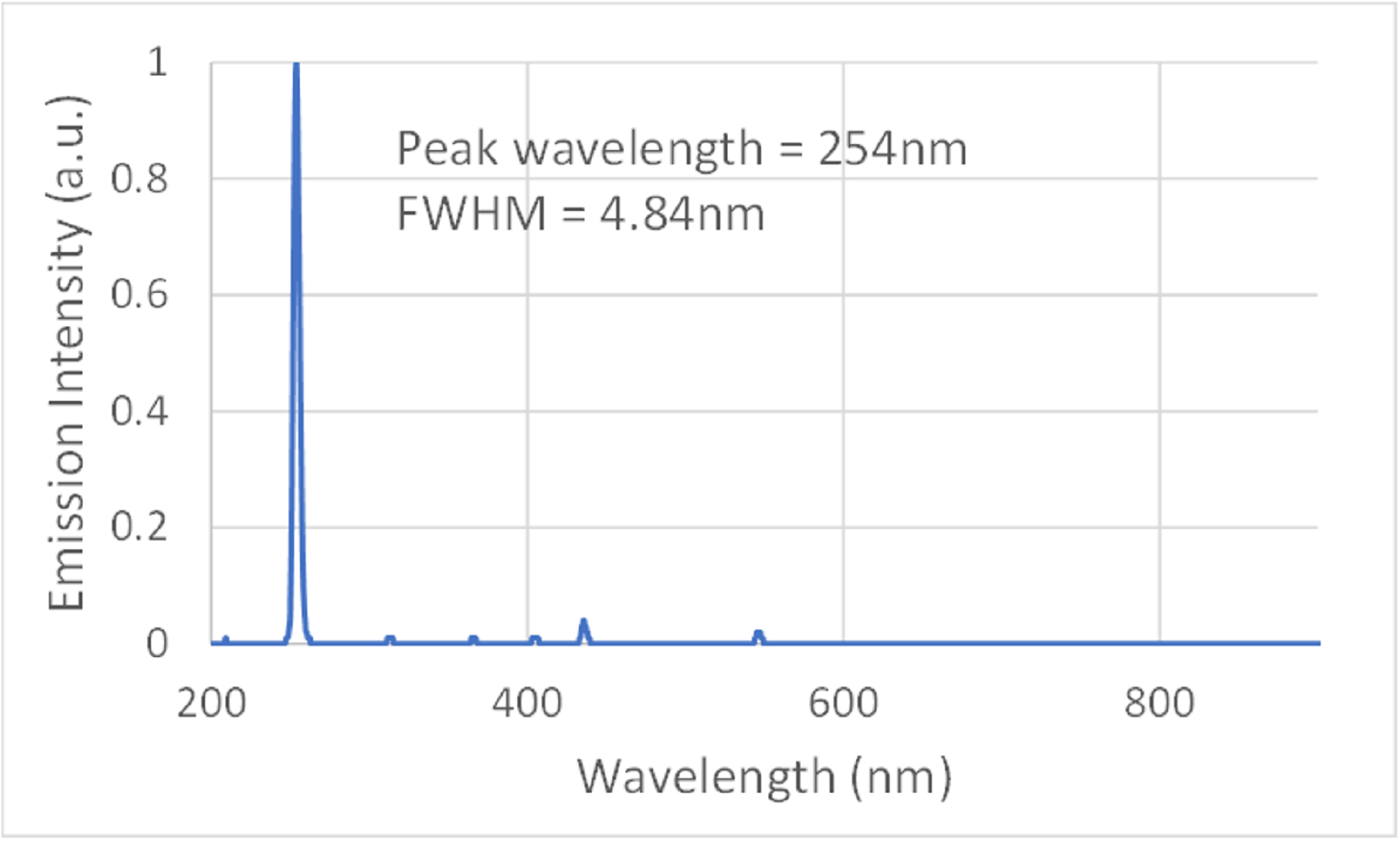
Spectrum of emission of the HNS 15 W G13 OSRAM lamp.

The first measurements were obtained with the grating located at a vertical distance of 10 cm from the lamps plane and with a single respirator inside the chamber located near the wall without lamps. The spectroradiometer (detector) was placed just at the side of the respirator closest to the wall without the lamp at the first measure (Figure 2a). An irradiance of 550 *μW/cm*^2^ was obtained. The measure was repeated moving the respirator 5 cm towards the opposite wall that contains a lamp (see Figure 2b). In this case an irradiance of 700 *μW/cm*^2^ was measured.

**Figure 2:**
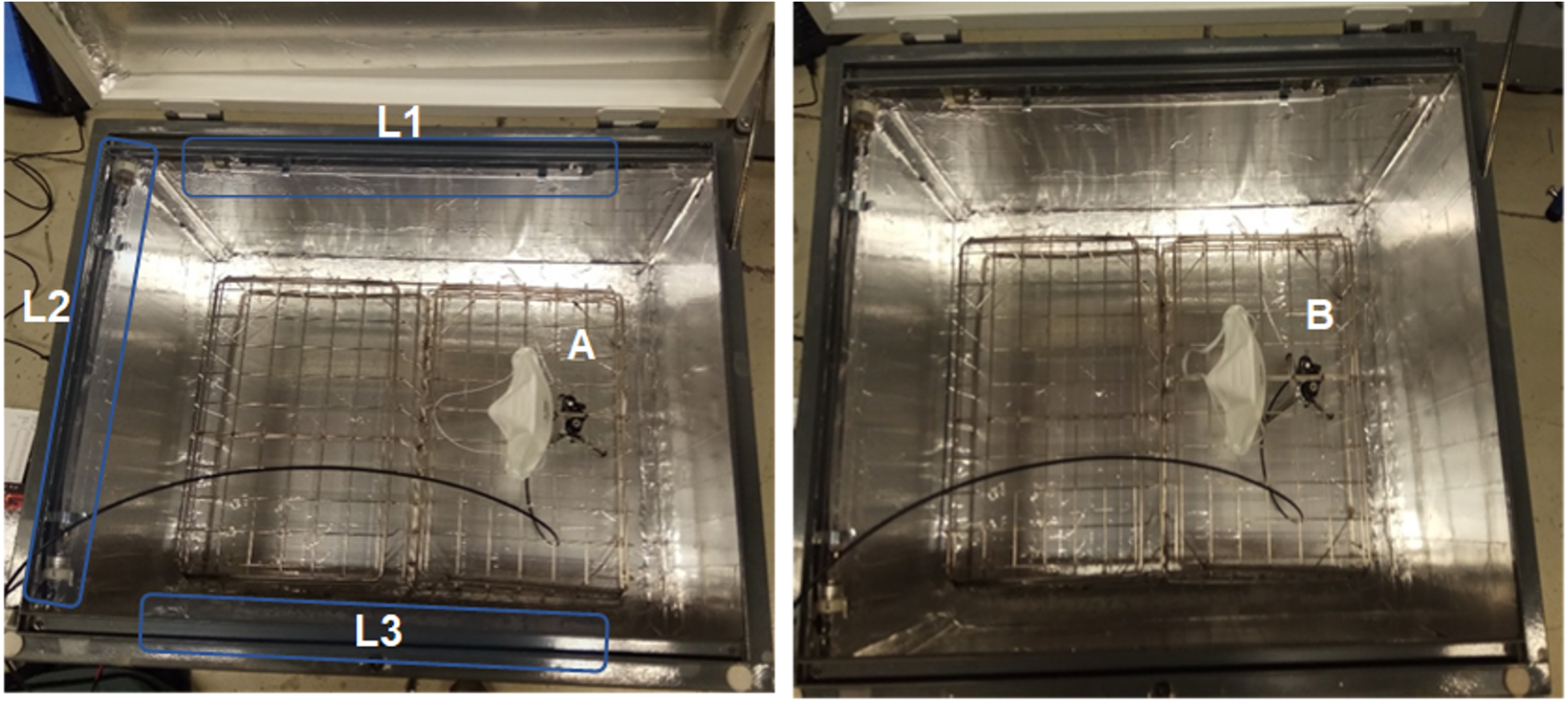
Photography of the inner part of the chamber, with the lamps L1 L2 and L3; the grating located at 10cm from the lamps plane; Spectrometer Position A, just beside to the right of the respirator (side closest to the wall without a lamp); Spectrometer Position B, with the respirator placed 5 cm towards the left side (towards lamp L2).

The same measurements were repeated with a distance between the grating and the lamps planes of 16 cm. In this case values of 650 *μW/cm*^2^ and 780 *μW/cm*^2^ were obtained at positions A and B, respectively. That indicates that a 16cm distance assures higher irradiances than at 10 cm distance, thus this height was selected for performing the following measurements. In addition, the detector was placed pointing upwards inside the masks, to measure the irradiance received by the inner part of the respirator. With this setup an irradiance of 60 *μW/cm*^2^ was obtained. (see Figure 3a). The first measurement over a facepiece respirator was done in position C shown in Figure 3b, that correspond to the position closer to the wall without lamp. Note that in this case the tallest model respirators are furthest away from L2. At this position, the detected irradiance was 470 *μW/cm*^2^, while at positions D and E the values obtained were 950 *μW/cm*^2^ and 1300 *μW/cm*^2^, respectively.

**Figure 3:**
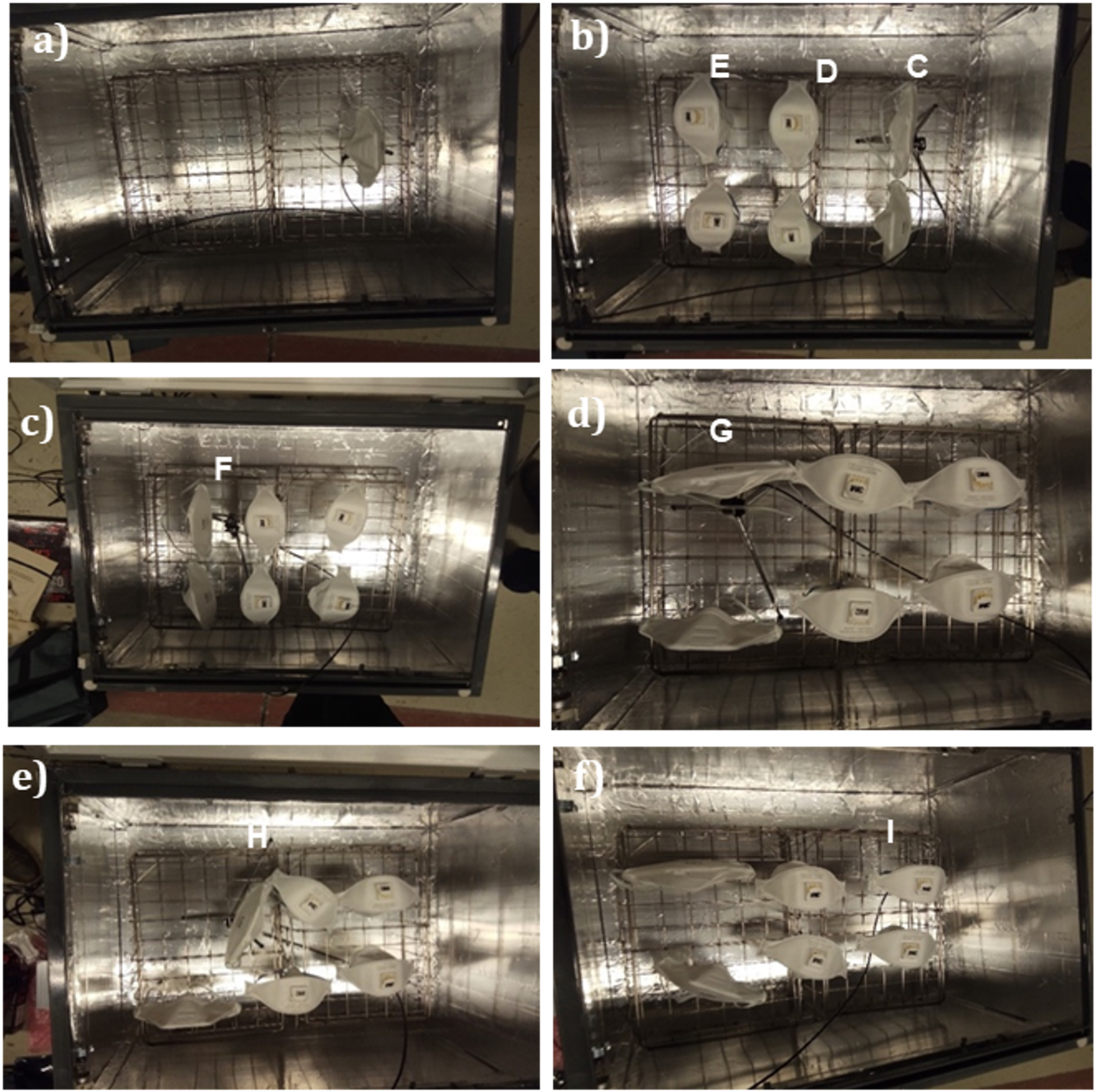
Photography of the inner part of the chamber, with the grating located at 16 cm from the lamps plane a) detector just below the respirator, and 6 respirators placed in different configurations: b) masks placed vertically with the shorter masks closer to the left lamp c) masks placed vertically with the taller masks closer to the left lamp d) masks placed horizontally, e) detector placed below one of the tall masks near the middle of the box; and f) detector placed below one of the short masks near the right side of the box, far from the lamp on the left side of the box.

In the second configuration, the position of the higher respirators was changed by moving them close to lamp L2. In this configuration, shown in Figure 3c, the irradiance achieved at the position marked by letter F was 1050 *μW/cm*^2^. To test if the irradiance depends on the position of the respirators over the grid, they were rotated an angle of 90 degrees and the sensor probe was bent at a 30 degree angle with the horizontal in order to evaluate the irradiance at the lateral of the respirators. This configuration is shown in Figure 3d. In this case the result obtained at the point marked by a G was 878 *μW/cm*^2^. Note that the sensor was located slightly below the plane of the masks so, a higher dosage value is expected in upper positions.

Finally, the sensor was placed under the respirators pointing downwards in order to determine the light coming from reflections at the bottom of the chamber at two different position marked by an H and an I in Figures 3 e and 3 f, respectively. The results at both positions were 422 *μW/cm*^2^ and 410 *μW/cm*^2^; respectively, indicating that the light distribution generated by reflections in the matte aluminum coating of the box is very homogeneous.

### Irradiance simulation

Figure 4 shows on the left column the experimental setup with the irradiance measured at different positions, and mask distribution. On the right column the results of the simulation are shown. The pictures present a pseudocolor map of the distribution of light inside the UVGI irradiator at the planes of the respiratory masks. Blue colors correspond to a reference amount of light. Green color represents a value equal to 3 times the reference value, yellow corresponds to 5 times, amber to 7 times, and red to 10 times. Similarity was observed between the measured data and the simulations. In both cases, as long as we move away from L2, a reduction in the irradiance is observed, getting the minimum exposure in the right side of the respirator mask on the right. Both, measured data and simulations reflect that in region C of Figure 4 we get half of the exposure obtained at D, and one third of that at E. Additionally, comparing the two light distribution obtained for the two orientations of the respiratory masks, the shadows obtained with the masks parallel to the long side of the box, are less pronounced (see the areas pointed by the arrows in Figure 4). Hence, this orientation is suggested as the preferred one.

**Figure 4:**
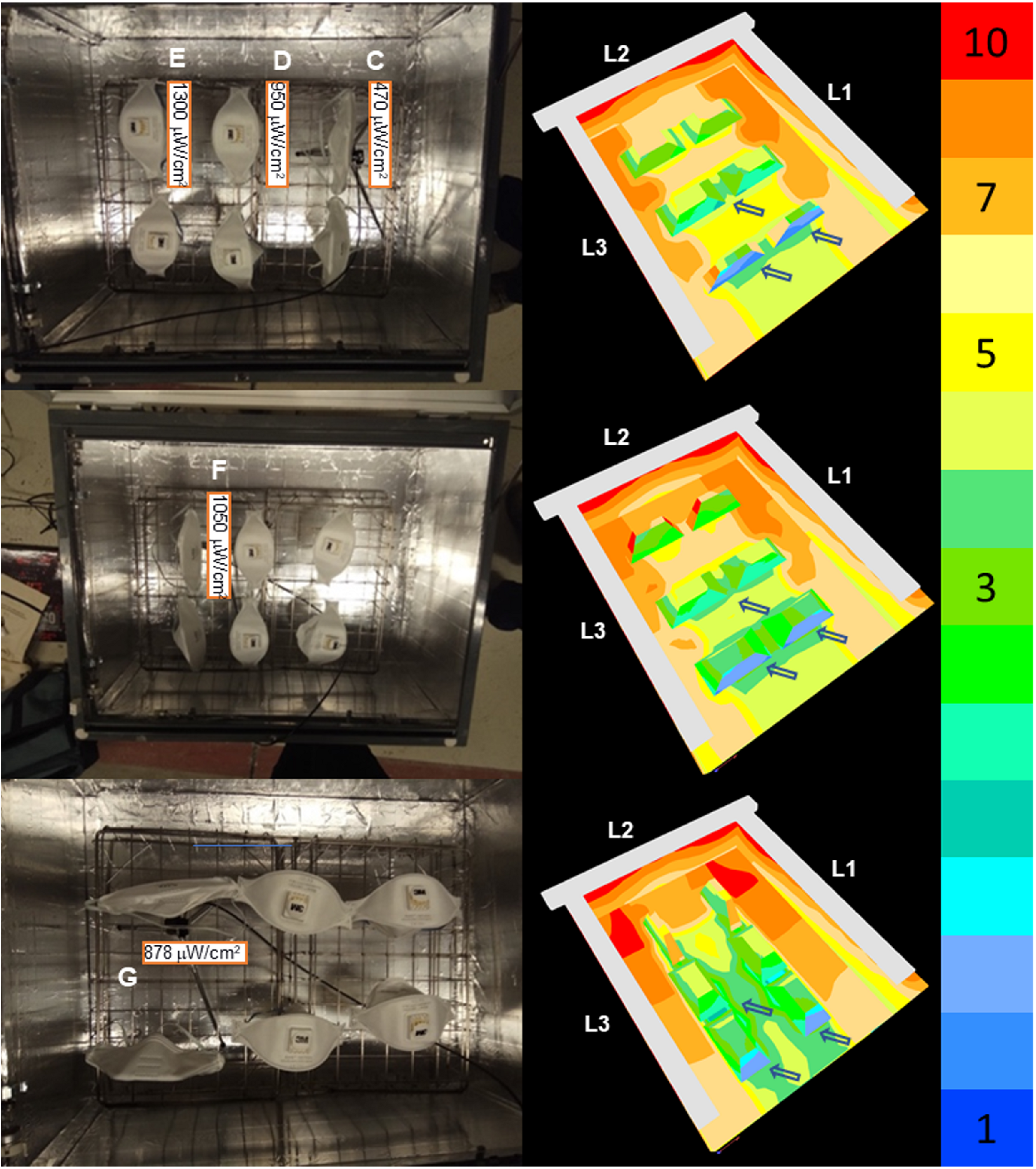
left) Representation of the experimental data obtained in the disinfection box; Right) simulated light distribution maps in pseudocolor maps inside the UVGI irradiator. The lamps are marked in white and named L1, L2 and L3. Blue colors correspond to a reference amount of light. Green color represents a value equal to three times the reference value, yellow corresponds to five times, amber to seven times, and red to 10 times.

Figure 5 compares the pseudocolor maps of the light distribution inside the disinfection box obtained by simulation, using three and four lamps. Note the difference between the two settings, as four lamps provide a more uniform light distribution with less difference in the light amount. With four lamps we also diminish the shadows in the face of the mask on the right side of the chamber.

**Figure 5:**
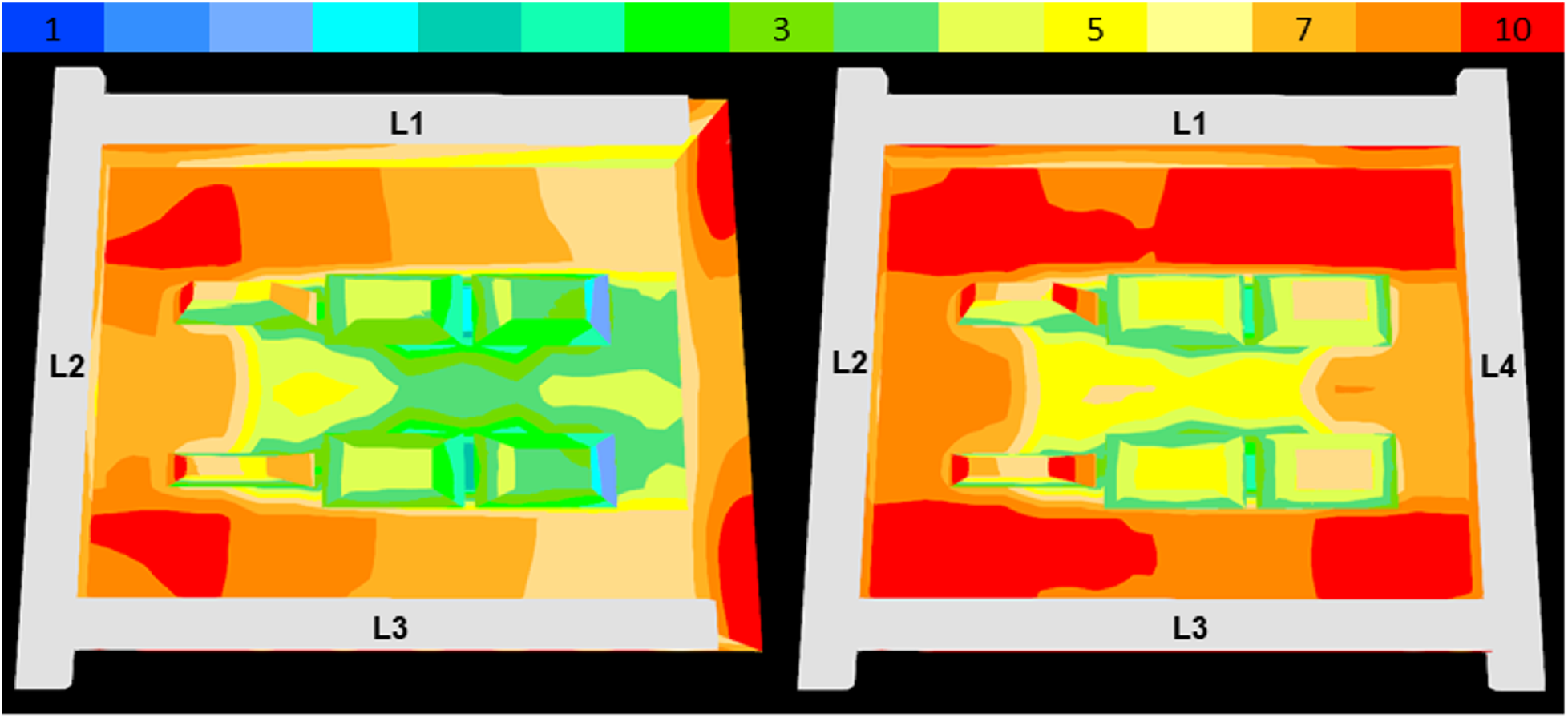
Pseudocolor map of light distribution obtained with three and four lamps for two models of facial respiratory masks, oriented parallel to the long side of the box.

### COVID-area application

Precise instructions were given for the UVGI irradiator use, indicating possible target times for decontamination based on current literature. Users were instructed to write down their name in either non-porous parts or in the elastic bands of their FFRs and mark them each time they underwent decontamination. The masks waiting to be decontaminated were placed inside individual, named cardboard envelopes by a staff worker who was protected by surgical mask, impermeable gown and gloves. The FFRs were then placed one by one as described in Fig 3d avoiding contact with the surface previously contacting the wearer and avoiding contact with the external UVGI box surface. After irradiation, masks were placed again in clean, individual envelopes. To ensure grid decontamination after each cycle, an additional time of 5 min was added with the cabinet closed and no masks inside. A warning card was placed in the box lid showing biohazard, UV-light hazard symbols, and a schematic representation of the mask placement (see Supplemental material). The names of the mask wearers, as well as the hour: minute irradiation intervals were written down in a list, next to the box.

## DISCUSSION

### Irradiance-angle relationship

This study demonstrates the extent of the dependence between dosimetry and mask location, relative to the light cone and other masks or obstacles that might be present. Regarding room UVC decontamination of FFRs, irradiance needs to be measured at the most extreme incidence angles. On the other side, small UVGI cabinets have less angle variability but they often need flipping the object to be decontaminated, and some respirator brands have conic-oval volumes that cannot stand stable in both flipped positions. A UVGI irradiation chamber is proposed which allows for multiple mask decontamination without the need to leave COVID-areas and does not require to flip the respirator for the desired dosage, thus ensuring minimal respirator manipulation. The data obtained reveal that the irradiance received inside the manufactured UVGI-box depends not only on the distance between the lamps plane and the base of the respirators but also, on the orientation and shape of the masks. This point becomes relevant to assure that all the respirators inside the chamber receive the correct dosage. Even though it could be expected that the nearer the respirator is to the lamps, the higher dose it receives, the experiment reflects this assumption is not true. For example, in the presented work (Figure 2a), 100 *μW/cm*^2^ more irradiance was obtained when placing the base of the masks at 16 cm than when placed at 10 cm from the plane that contains the lamps.

The absorption produced in a respirator can also be determined by measuring the dosage received immediately below one of them. The data confirms that around one order of magnitude of the dosage is absorbed by the bulk of the respirator. This should be taken into account in case that the geometry of the respirator does not allow to flip it to receive a certain dose for sterilization. In this case, the exposure time should be calculated to warrant the dosage in the inner part of the FFRs.

### Dosage-time relationship

The resulting UV dose (fluence) is the product of the irradiance by exposure time, as follows:

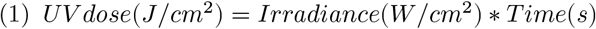

Therefore, in order to know the exposure time needed for sterilization of the respirators used with COVID patients, the following relation is used:

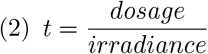

being **t**, the exposure time expressed in seconds, **dosage** expressed in *J/cm*^2^ and the **irradiance** in *W/cm*^2^.

The time needed to receive a range of different possible dosages in the less irradiated position was calculated. As an example, exposure times for different dosages are presented at Table 1.

**Table 1.** Exposure time with the respirators in the configuration of **Fig 3d**.

| **Dosage** | **5mJ/cm2** | **300mJ/cm2** | **1J/cm2** | **3J/ cm2** |
| --- | --- | --- | --- | --- |
| Exposure time | 11 s | 12 min | 36 min | 1 h 40 min |

This was carried out to ensure that the cumulative dosage received by all and each respirator is above the minimum dosage determined by the user in order to feel safe in reusing the masks, after being in contact with COVID patients.

### UVGI disinfection

Ultraviolet light is gaining acceptance among the healthcare community as they are a cost-effective alternative to heat or chemical decontamination. At moderate UVGI doses, mask performance still surpasses that of surgical masks, thus being a viable alternative when no new FFRs are available. Viscusi et al administered 3.24 *J/cm*^2^ and examined fit, odor, comfort and deterioration in several mask brands, not finding significant differences using UVGI (Viscusi et al., 2011). NIOSH collaborators, Lindsley et al (Lindsley et al., 2015) found changes in particle penetration, but only small changes in resistance after very high UVGI doses (up to 950 *J/cm*^2^).

Regarding disinfection, NIOSH guidelines (Lindsley et al., 2015) advise to discard masks after aerosol-generating procedures. However, previous studies have shown that UV disinfection is suitable to remove viral load although more studies are needed to ascertain viral removal from the inner FFR layers. UVGI dose for coronavirus in surfaces has been shown to be lower than other types as they are single-stranded RNA virus. For example, Duan et al (Duan et al., 2003) found that 0.32 *J/cm*^2^ can inactivate SARS-CoV in culture plates, whereas for H1N1 influenza, decontamination with 1.2, 1.8 or 1.98 Joules/cm^{2} achieved an average 4-log reduction of viable H1N1 influenza virus (Heimbuch et al., 2011; Lore et al., 2012; Mills et al., 2018). In our COVID ICU, we chose a target dosage of 3 *J/cm*^2^ because it is above to the lethal dose of influenza virus (1.5-2 *J/cm*^2^), given that the coronavirus peak overlapped with an influenza peak during March-April in our region.

Several institutions such as NIOSH *(CDC - Recommended Guidance for Extended Use and Limited Reuse of N95 Filtering Facepiece Respirators in Healthcare Settings - NIOSH Workplace Safety and Health Topic*, 2020) or the ECDC (“Cloth Masks and Mask Sterilisation as Options in Case of Shortage of Surgical Masks and Respirators,” 2020), as well as the mask manufacturer 3M (3MCompany, 2020) discourage FFR reuse except in extreme shortages when no new masks are available. Based on recommendations given by those sources and our own user experience, an additional set of instructions were given to promote a rational use of the irradiator when caring for COVID patients. UVGI irradiator use was discouraged when any of the following conditions are met:

1- The specified time of usage has been completed for one particular mask (e.g. 8h total use for N95 masks).
2- FFR which completed 3 UV-C cycles (equivalent to 9 *J/cm*^2^ in total).
3- Used during aerosol-generating procedures (such as oral hygiene or airway procedures).
4- When an FFR is contaminated with patient fluids.
5- When an FFR is wet (sweat, etc.).
6- When any of the 3 known complications UV-C disinfection in FFRs:
6a- Loss of fit or adjustment after the user seal check.
6b- Moderate or intense odor that doesn’t disappear after 10 minutes of aireation.
6c- Elastic bands deterioration. The stapling of replacement bands has to ensure that the perforations are covered
7- Also, discard when any FFR:
7a- Hasn’t undergone adequate decontamination in a suitable time.
7b- Has visible damage to the mask or increased difficulty in breathing through the filter (upon user seal check).

### Limitations

This is a study where changes in irradiance are studied in a closed, controlled environment. Different mask brands have different shapes, modifying local irradiance. To compensate for this, overdosing of more exposed areas might be necessary, causing them to accumulate more deterioration, shortening the respirator’s life. Further studies might be needed to ascertain dose homogeneity when the UVGI lamps are placed in a bigger compartment, such as a room. In addition, only a 3-lamp setting was tested and used due to space limitations in the remaining side, between the box closing mechanism and the grate. UV-transparent quartz lamps have a higher risk of breakage, exposing the users to toxic mercury vapor. Therefore, due to safety concerns we decided not to place a fourth lamp in the first UVGI box. Adding a fourth lamp or light sources on both sides of the rack might provide more reliable illumination, avoiding both over and underexposure. This UVGI box currently does not support the decontamination of more than six masks at once. Bigger designs can provide mask reuse at a bigger scale in times of severe shortages. The impact of treatment on filter penetration was not assessed in this study. Respirator seal, fit or comfort were only checked by the mask user. Currently, virologic assessment is being designed in an appropriate setting for this irradiator. Thus, explored dosage range regimes are based upon previously published experiments elsewhere.

## CONCLUSION

Irradiance over FFR surfaces depend on several factors such as distance and angle of incidence of the light source. Careful irradiance measurement and simulation can ensure reliable dosage in the whole mask surface, balancing overexposure. Closed box systems might provide a more reliable, reproducible UVGI dosage than open settings.

## RECOMMENDATIONS

- Custom UVGI devices must feature mechanisms to protect from harmful UVGI irradiation.
- Dosimetry from strategic locations of an UVGI facility allows for correct time-irradiance calculations in respirators at different positions.
- Irradiance measurements can be performed by experts in visible light pollution or photonics, given access to a UV-C light spectroradiometer.
- Alternatively, manual dosimeter probes can be used at such locations.
- Careful respirator placement must be ensured to minimize error in the administered UV dose.
- Clear instructions on device operation and respirator reuse must be issued, updated and published in the work environment.
- Careful user seal checks must be performed after reuse of a decontaminated mask.

## Data Availability

Available data are included in the manuscript

## Acknowledgements

We would like to acknowledge the help and support received by the Physics and Optics faculties, as well as to Ricardo Rodríguez. We would also like to thank Professor Salvador Bará Viñas of the Photonics4Life group at USC for his advice in carrying out measurements and checking calculations.

## Disclosures

Authors declare that there are not conflicts of interest related to the results of this article.

## Funding

This work was partially supported by the Consellería de Educación Program for Development of a Strategic Grouping in Materials - AeMAT Grant No. ED431E2018/08 and Xunta de Galicia ref. ED431B2017/64.

**Abbreviations**
CDC: Centers for Disease Control and Prevention.
CoV2: novel coronavirus 2.
FFR: filtering facepiece respirator.
NIOSH: National Institute for Occupational Safety and Health.
SARS: severe acure respiratory syndrome.
UV-C: ultraviolet C.
UVGI: ultraviolet germicidal irradiation.

## Notes

### Competing Interest Statement

The authors have declared no competing interest.

### Funding Statement

This work was partially supported by the Conselleria de Educacion Program for Development of a Strategic Grouping in Materials - AeMAT Grant No. ED431E2018/08 and Xunta de Galicia ref. ED431B2017/64.

### Summary of Updates

Updated preprint structure to improve legibility

## REFERENCES

3MCompany. (2020). Decontamination Methods for 3M Filtering Facepiece Respirators such as N95 Respirators. 3M Technical Bulletin, *Revision 6*.

Bass, M., DeCusatis, C., Enoch, J., Lakshminarayanan, V., Li, G., MacDonald, C., Mahajan, V., & Stryland, E. V. (2009). Handbook of Optics, Third Edition Volume IV: Optical Properties of Materials, Nonlinear Optics, Quantum Optics (3 edition). McGraw-Hill Education.

CDC. (2020). Coronavirus Disease 2019 (COVID-19). In Centers for Disease Control and Prevention. https://www.cdc.gov/coronavirus/2019-ncov/hcp/respirators-strategy/crisis-alternate-strategies.html

CDC - Recommended Guidance for Extended Use and Limited Reuse of N95 Filtering Facepiece Respirators in Healthcare Settings - NIOSH Workplace Safety and Health Topic. (2020). https://www.cdc.gov/niosh/topics/hcwcontrols/recommendedguidanceextuse.html

Cloth masks and mask sterilisation as options in case of shortage of surgical masks and respirators. (2020). In European Centre for Disease Prevention and Control. https://www.ecdc.europa.eu/en/publications-data/cloth-masks-sterilisation-options-shortage-surgical-masks-respirators

Duan, S.-M., Zhao, X.-S., Wen, R.-F., Huang, J.-J., Pi, G.-H., Zhang, S.-X., Han, J., Bi, S.-L., Ruan, L., & Dong, X.-P. (2003). Stability of SARS Coronavirus in Human Specimens and Environment and Its Sensitivity to Heating and UV Irradiation. Biomedical and Environmental Sciences: BES, 16, 246-255.

Heimbuch, B. K., Wallace, W. H., Kinney, K., Lumley, A. E., Wu, C.-Y., Woo, M.-H., & Wander, J. D. (2011). A pandemic influenza preparedness study: Use of energetic methods to decontaminate filtering facepiece respirators contaminated with H1N1 aerosols and droplets. American Journal of Infection Control, 39(1), e1-9. https://doi.org/10.1016/j.ajic.2010.07.004

Lindsley, W. G., Jr, S. B. M., Thewlis, R. E., Sarkisian, K., Nwoko, J. O., Mead, K. R., & Noti, J. D. (2015). Effects of Ultraviolet Germicidal Irradiation (UVGI) on N95 Respirator Filtration Performance and Structural Integrity. Journal of Occupational and Environmental Hygiene, 12(8), 509-517. https://doi.org/10.1080/15459624.2015.1018518

Lore, M. B., Heimbuch, B. K., Brown, T. L., Wander, J. D., & Hinrichs, S. H. (2012). Effectiveness of three decontamination treatments against influenza virus applied to filtering facepiece respirators. The Annals of Occupational Hygiene, 56(1), 92-101. https://doi.org/10.1093/annhyg/mer054

Mills, D., Harnish, D. A., Lawrence, C., Sandoval-Powers, M., & Heimbuch, B. K. (2018). Ultraviolet germicidal irradiation of influenza-contaminated N95 filtering facepiece respirators. American Journal of Infection Control, 46(7), e49-e55. https://doi.org/10.1016/j.ajic.2018.02.018

Viscusi, D. J., Bergman, M. S., Novak, D. A., Faulkner, K. A., Palmiero, A., Powell, J., & Shaffer, R. E. (2011). Impact of three biological decontamination methods on filtering facepiece respirator fit, odor, comfort, and donning ease. Journal of Occupational and Environmental Hygiene, 8(7), 426-436. https://doi.org/10.1080/15459624.2011.585927

Welch, D., Buonanno, M., Grilj, V., Shuryak, I., Crickmore, C., Bigelow, A. W., Randers-Pehrson, G., Johnson, G. W., & Brenner, D. J. (2018). Far-UVC light: A new tool to control the spread of airborne-mediated microbial diseases. Scientific Reports, 8(1), 1-7. https://doi.org/10.1038/s41598-018-21058-w

